# Determining the Relationship between Seizure-Free Days and Other Predictors of Quality of Life in Patients with Dravet Syndrome and Their Carers

**DOI:** 10.1101/2022.10.07.22280500

**Authors:** A Pinsent, G Weston, EJ Adams, W Linley, N Hawkins, M Schwenkglenks, C Hamlyn-Williams, T Toward

## Abstract

**Objectives:** Dravet syndrome (DS) is a rare, lifelong epileptic encephalopathy characterised by frequent and severe seizures associated with premature mortality. Typically diagnosed in infancy, patients also experience progressive behavioural, motor-function and cognitive decline. Twenty percent of patients do not reach adulthood. Quality of life (QoL) is impaired for both patients and their carers. Reducing convulsive seizure frequency, increasing seizure free days (SFDs) and improving patient/carer QoL are primary treatment goals in DS. This study explored the relationship between SFDs and patients’ and carers’ QoL to inform a cost-utility analysis of fenfluramine.

**Methods:** In fenfluramine registration studies, patients (or their carer proxies) completed the Paediatric Quality of Life inventory (PedsQOL). These data were mapped to EuroQol-5 Dimensions Youth version (EQ-5D-Y) to provide patient utilities. Carer utilities were collected using EQ-5D-5L and mapped to EQ-5D-3L to align patient and carer QoL on the same scale. Linear mixed-effects and panel regression models were tested and Hausman tests identified the most appropriate approach for each group. On this basis, a linear mixed-effects regression model was used to examine the relationships between patient EQ-5D-Y, and clinically relevant variables (age, frequency of SFDs per 28-days, motor impairments and treatment dose). A linear panel regression model examined the relationship between SFDs and carer QoL.

**Results:** Adjusting for age and underlying comorbidities, the patient regression model showed that SFDs per 28-days was a significant predictor of QoL. Each additional patient-SFD increased utility by 0.005 (*p*<.001). The carer linear panel model also showed that increasing SFDs per 28-days was a significant predictor of improved QoL. Each additional SFD increased carer utility by 0.014 (*p*<.001).

**Significance:** This regression framework highlights that SFDs are significantly correlated with both patients’ and carers’ QoL. Treatment with effective antiseizure medications that increase SFDs, directly improves QoL for patients and their carers.

**Short summary:** DS patients experience daily severe seizures with progressive deterioration in their physical, cognitive and behavioural development (“comorbidities”), which substantially impacts the QoL of patients and their carers. Reducing seizure frequency and increasing Seizure Free Days (SFDs) are key treatment goals. This study examined the relationship between seizures and patients’ and carers’ QoL. Regression analyses were conducted using data from the fenfluramine registration studies and confirmed increasing SFDs directly, and quantifiably, improved patient and carer QoL (utilities per additional SFD per 28-days: patient=0.005 and carer=0.014). These analyses demonstrate that effective antiseizure treatment can directly and profoundly improve patients’ and carers’ QoL.

## Introduction

Dravet syndrome (DS) is a rare and lifelong epileptic encephalopathy, characterised by frequent and often severe seizures that are resistant to treatment with existing antiepileptic drugs (AEDs); sustained seizure freedom is rarely achieved (1,2). In addition to the risks of premature mortality due to sudden unexpected death in epilepsy (SUDEP), status epilepticus and accidents (3,4), high convulsive seizure frequency in DS is associated with an early onset of progressive comorbidities such as neurodevelopmental and motor impairments, and behavioural difficulties (5), which have implications for independent living (6,7). The combination of often daily seizures, and cognitive, motor, behavioural and sleep impairments, significantly impairs the quality of life (QoL) of DS patients (5), and can exert a substantial burden on families, with parents and carers of patients often giving up paid employment to be full-time caregivers with little respite from their carer responsibilities (8–11). Patient groups have therefore reported that DS has a profound impact on both patients’ and their caregivers’ QoL (8).

Fenfluramine (FFA) is a recently licensed add-on treatment option for patients with DS. Two registration, phase III randomised, placebo-controlled trials (Study 1 (12) and Study 2 [also known as Study 1504] (13)) have shown fenfluramine, when added to current standard of care AEDs, profoundly reduces convulsive seizures and provides a sustained and durable response over at least 3 years of observation. Uniquely, these studies also assessed QoL of both patients and their carers. To inform a health technology assessment (HTA) of FFA by the National Institute for Health and Care Excellence (NICE) in the UK (14), the current study used patient-level data from these trials to explore the relationship between seizures and QoL for patients and carers.

A systematic literature review (SLR) of QoL studies was undertaken as part of the NICE appraisal process and highlighted a substantial impact of DS on both patients’ and carers’ QoL (14). Several studies reported a reduced QoL with increasing seizure frequency. However, the magnitude of this relationship was not quantified (15,16), and there was a paucity of research exploring whether short-term periods of seizure freedom (rather than longer-term remission) or complete seizure freedom (rarely obtained in DS), may be an important and meaningful metric in quantifying the burden of illness. To help address this gap, a recent study with DS expert clinicians was conducted (17), and confirmed that a convulsive seizure-free day (SFD) was directly relevant to patients’ and carer’s QoL. Increasing the number of convulsive SFDs is therefore also expected to reduce the physical burden, anxiety and fears experienced by caregivers, and improve their QoL. Thus, alongside reduced convulsive seizure frequency, increased SFDs is considered a key therapeutic goal for patients with DS and their carers.

This study aimed to quantify the impact of SFDs and other clinical covariates to understand which factors may predict DS patient and carer QoL. Furthermore, to support the use of these data for cost-effectiveness analyses that incorporate quality-adjusted life years (QALY), the QoL measures were transformed to utility values.

## Methods

Study 1 (12) and Study 2 (13) evaluated the efficacy and safety of FFA as an as an add-on therapy to standard of care AEDs for the treatment of seizures associated with DS. In pharmacokinetic and pharmacodynamic studies, an interaction with stiripentol was identified, and a bioequivalent dose of FFA when used with concomitant stiripentol was determined. Study 1 investigated FFA (0.7mg/kg/day, up to a maximum daily dose of 26mg) or placebo, when added to patient’s existing standard of care AEDs that excluded stiripentol. Study 2 investigated FFA (0.4mg/kg/day, up to a daily maximum dose of 17mg) or placebo, when added to patient’s existing standard of care that included stiripentol. Other than a respective 2- and 3-week titration period for FFA (or placebo), the trials were of similar design and conducted in similar geographies at the same time. QoL was assessed in both patients and carers in both trials. In initially reported analyses of the FFA registration trials, no adjustment of QoL for covariates related to SFDs was undertaken.

The purpose of this study was to assess the impact of SFDs on patient and carer QoL, adjusting for significant covariates, and to derive utility scores associated with the changes in SFD observed with treatment in the FFA trials.

## Measures

### Paediatric Quality of Life Inventory (PedsQL)

In both of the FFA registration trials (12,13), Paediatric QoL was assessed using the PedsQL Version 4 Generic Score Scale (18). This consists of four domains measuring physical, emotional, social, and school functioning. The scale is available in age-appropriate formats, with child self-report and parent proxy-report formats. In the trials, the age-appropriate instruments used were (for ages 2-4, 5-7, 8-12, and 13-18 years), in addition to the parent instrument. PedsQL was completed by patients or their proxies. Scores were expressed on a scale of 0-100, in which higher scores represented better QoL.

### EuroQol -5 Dimensions five-level (EQ-5D-5L)

Caregiver QoL was assessed using the EQ-5D-5L instrument (19). The measure comprises five dimensions: mobility, self-care, usual activities, pain/discomfort, and anxiety/depression. A five-level rating was given for each dimension. A score is expressed on a scale of 0-100, with 100 being the best health you can imagine, and 0 equating to the worst health that can be imagined by the respondent.

### Data on clinically relevant variables

Individual-level data for patients and carers were obtained from the FFA trials (12,13), including: patient age group, treatment group (placebo or treatment), study cohort, visit number, presence of motor impairments, 28-day frequency of SFDs.

### Motor impairments

Individuals were assigned motor impairment categories (none, ataxia, and severe) based on the medical history terms provided within the FFA registration trials. Individuals were assigned to each group if it was reported that they had the following:

- None: no ataxia, gait, hypotonic, motor, ambulatory, wheelchair or such keywords,
- Ataxia: ataxia, gait
- Severe: profound, severe, acute, wheelchair, non-ambulatory, cerebral palsy.

### 28-day frequency of SFDs

During clinical trials (12,13) seizure frequency and SFDs were recorded at baseline and throughout the trials using electronic diaries completed by carers on a daily basis. A patient was considered as having a seizure day if they had at least 1 convulsive seizure that day. Any day with no convulsive seizures was considered a SFD.

#### Collection and mapping of patient and carer utilities

PedsQL data were collected from 155 patients (52 in Study 1 and 76 in Study 2), or their proxy (who was the same throughout the study), at three-time points in the FFA registration studies (20,21); week 6 (randomisation to treatment initiation, visit 1), week 12 (after end of titration period of 2-3 weeks depending on study, visit 2), and week 20 (end of maintenance period or discontinuation, visit 3) – hereafter referred to as visits. Complete PedsQL data for all visits were available for 128/155 patients.

To obtain patient EQ-5D utility values, a widely accepted measure of QoL and are typically used in cost-utility analyses, the PedsQL data were mapped directly to EQ-5D-Y using the Khan *et al* UK-based algorithm (22) which is the only published and validated mapping algorithm available to estimate patient utilities from PedsQL data.

EQ-5D-5L data were collected directly from 185 carers in the registration studies at two time points: visit 1 and visit 3. Data for 176 carers (106 in Study 1 and 70 in Study 2) were available for both visits. As per the 2019 NICE position statement (23), all data were mapped from EQ-5D-5L onto EQ-5D-3L using the UK value set developed by van Hout *et al* (24). To enable a comparative assessment of the relationship between the carer’s and patient’s QoL; as well as to derive a common utility measurement for an economic evaluation that utilised a QALY metric it was important to have the same scaling for both patients and carers. A transformation of the carer EQ-5D-5L data was therefore undertaken to derive EQ-5D-3L values, which is comparable to the patient EQ-5D-Y.

EQ-5D data for both patients and carers were multiplied by 100 to achieve a 0-100 scale.

EQ-5D utilities and PedsQL values were evaluated as the outcome variables of the present analysis for all individuals for which complete case data was available. Complete case data was needed as the regression framework was not able to accommodate missing data for the outcome variables.

## Analysis

### Statistical analysis of patient and carer data

Whilst fixed effects models are considered to be the gold standard for data structures like the one under study, it is not always possible to fit all effects reliably to all patients (25). Thus, a more parsimonious mixed-effects approach may be better suited to such data. Hence, two types of regression models were considered after the initial data description and applied to each dataset;

1. linear mixed-effects regression models. Given that repeated measures were taken for each patient at 2 or 3 different points in the trial, as categorical variables both subject ID and visit numbers were considered as random effects in the models. Data for both patients and carers were then analysed separately to assess whether there were any differences in QoL scores between patients themselves and between visits;
2. panel linear fixed-effects regression models. Variables that change little, or not at all, over time should not be included in such models because they produce collinearity with the fixed effects. Therefore, only covariates which varied over time were considered.

We then assessed which of these two model types represented the data better. To determine which model was more appropriate for the data the final linear mixed-effects models for patients and carers were statistically compared to the final panel linear fixed-effects models for the same datasets, using the Hausman test (26),. The model framework that was statistically supported by the Hausman test was taken forward as the final model for the QoL datasets for patients and carers.

All analyses were conducted in R (R core Team, 2019). The function Imer from the package Ime4 (27) was used to produce the linear mixed-effects models; confidence intervals and *p-values* were constructed using degrees of freedom from Kenward-Roger’s method (28). The plm function was used to generate the panel linear fixed-effects models. Plots were produced with ggplot2 (29)) and tables with sjPlot (30).

### Covariate selection

The initial selection of covariates from the clinical trial data (12,13) was based on availability of data collected, clinical expert advice and guidance from the literature obtained through the initial SLR (31). Covariates explored for both the carer and patient models were:

- Motor impairments (none, ataxia, or severe)
- Visit number during the trial period
- 28-day frequency of SFDs
- RCT (trial) (Study 1 or Study 2)
- Patient age group

These clinical covariates were selected as candidate predictors for the linear mixed-effects models based on their ability to predict EQ-5D-Y and PedsQL scores of patients and EQ-5D-3L scores of carers in univariate analysis. Decision criteria included statistical significance (assessed by a two-sided *p-value* <0.05) in the baseline data and/or clinical understanding of their relevance to QOL outcomes. The study arm within the trial was not included as a covariate due to its substantial correlation with SFDs. The univariate analysis was conducted using the baseline EQ-5D-Y data and PedsQL data for patients, and EQ-5D-3L data for carers as the outcome variables. Following the univariate analysis, interaction terms between all covariates were tested using the baseline data for the: patient PedsQL data, the transformed patient EQ-5D-Y and carer EQ-5D-3L data. If an interaction was found to be significant it was tested again following the selection of the main effects in the multivariate regression model. Forward selection of main effects for the linear mixed effects model was conducted using patient and carer QoL data measured at all time points to determine the fixed effects in the final model. Statistically significant covariates (*p* <0.05) were retained in the final models.

In the panel linear model analysis, 28-day frequency of SFDs was tested as a covariate. No other variables (i.e. patient age group, study, or motor impairments) in the data varied over time.

### Predicting utility scores

With the estimated relationship between QoL outcome data for patients and carers and clinically relevant covariates calculated through the final models, patient and carer utility scores were predicted for each SFD. Marginal means were computed with Ismeans (32) and the predicted relationship was plotted to visually assess the relationship between SFDs and patient and carer QoL.

## Results

In the reported patient level data in Table 1, there was a slight increase in the mean unadjusted PedsQL score in Study 1 for patients on treatment. Similarly, for carers, there was an increase in the mean EQ-5D-3L score in the treatment arms of both studies. The mean number of SFDs experienced by patients in both treatment groups increased during the trial, whilst those in the placebo arms remained consistent throughout the trial period.

**Table 1.**
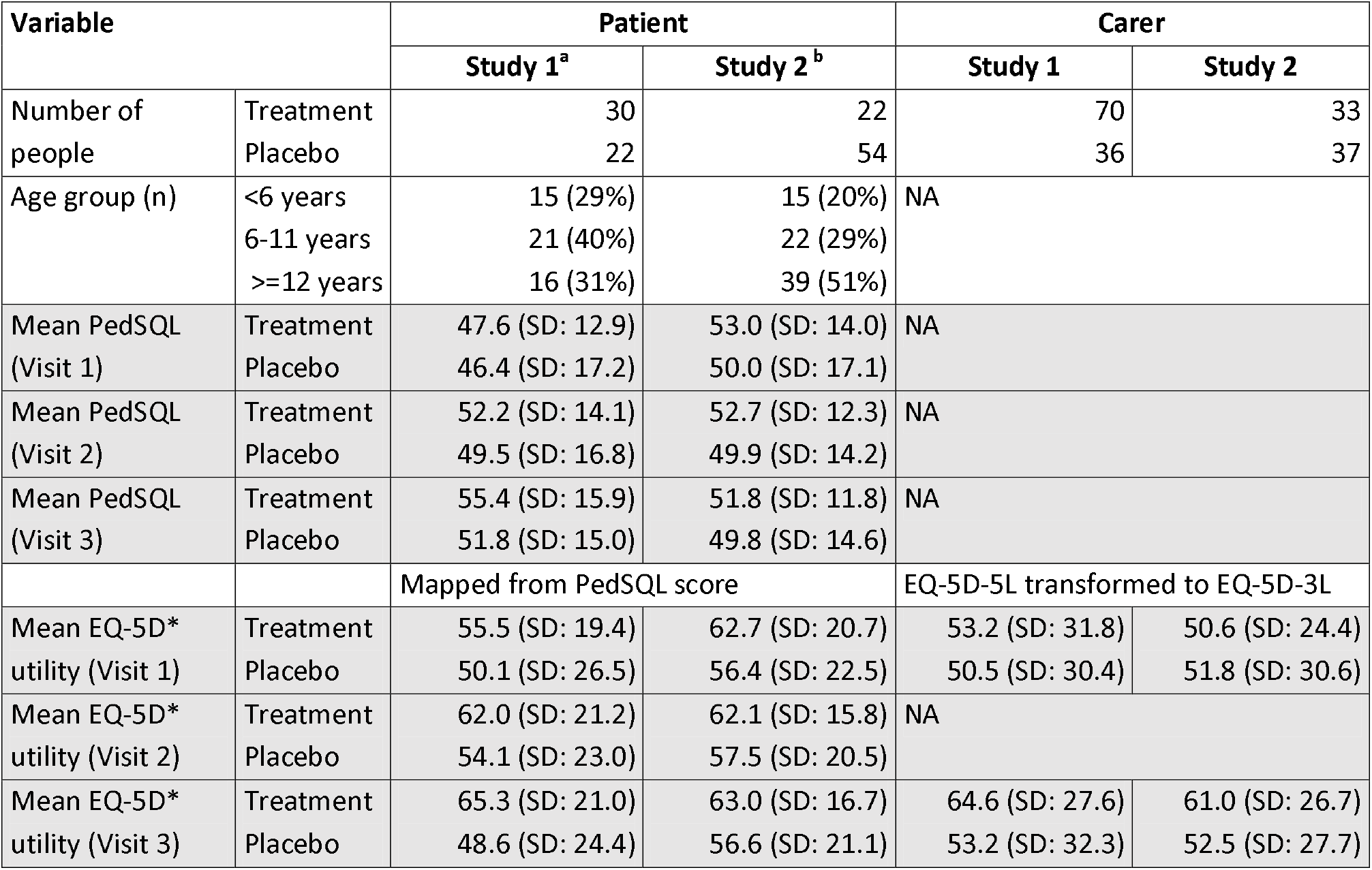
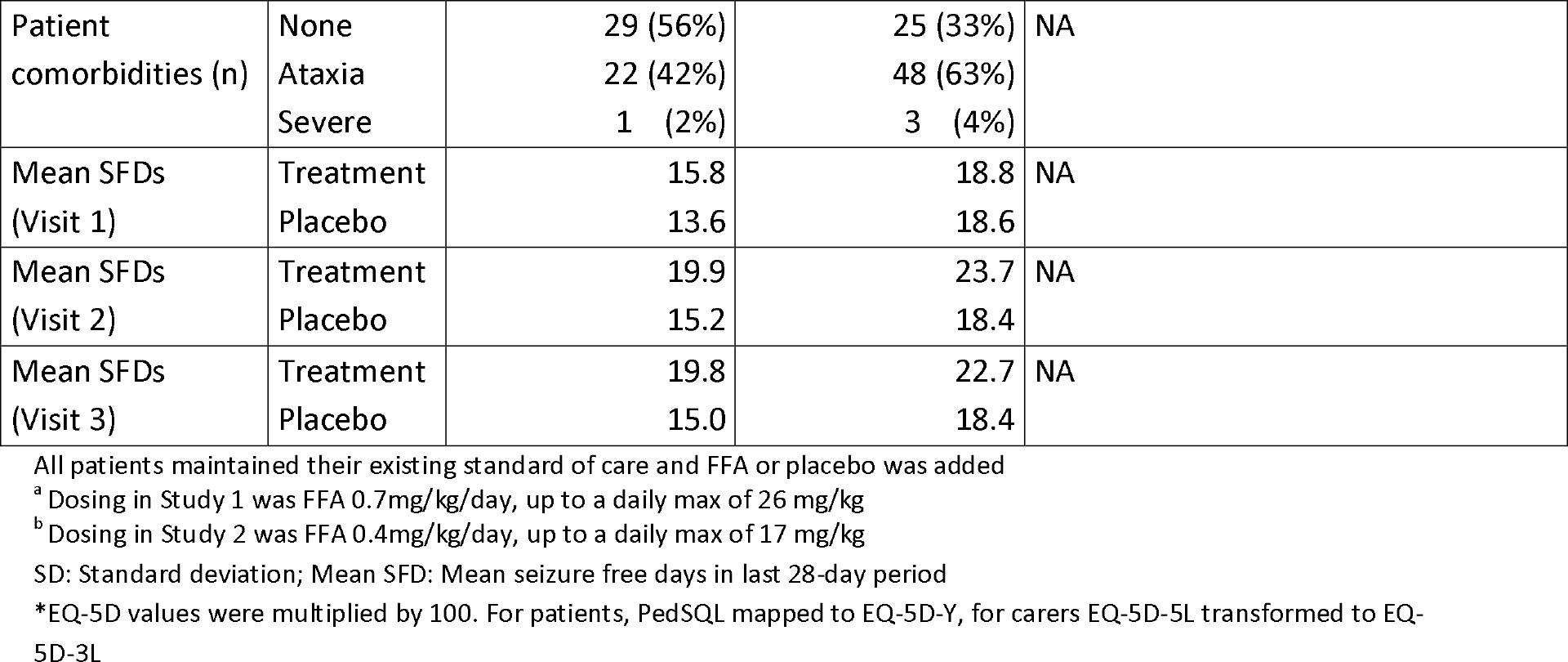
Summary of the unadjusted patient and carer data collected in the Fenfluramine registration RCTs.

| Variable |  | Patient |  | Carer |  |
| --- | --- | --- | --- | --- | --- |
|  |  | Study 1<sup>a</sup> | Study 2<sup>b</sup> | Study 1 | Study 2 |
| Number of people | Treatment | 30 | 22 | 70 | 33 |
|  | Placebo | 22 | 54 | 36 | 37 |
| Age group (n) | <6 years | 15 (29%) | 15 (20%) | NA |  |
|  | 6-11 years | 21 (40%) | 22 (29%) |  |  |
|  | >=12 years | 16 (31%) | 39 (51%) |  |  |
| Mean PedSQL (Visit 1) | Treatment | 47.6 (SD: 12.9) | 53.0 (SD: 14.0) | NA |  |
|  | Placebo | 46.4 (SD: 17.2) | 50.0 (SD: 17.1) |  |  |
| Mean PedSQL (Visit 2) | Treatment | 52.2 (SD: 14.1) | 52.7 (SD: 12.3) | NA |  |
|  | Placebo | 49.5 (SD: 16.8) | 49.9 (SD: 14.2) |  |  |
| Mean PedSQL (Visit 3) | Treatment | 55.4 (SD: 15.9) | 51.8 (SD: 11.8) | NA |  |
|  | Placebo | 51.8 (SD: 15.0) | 49.8 (SD: 14.6) |  |  |
|  |  | Mapped from PedSQL score |  | EQ-5D-5L transformed to EQ-5D-3L |  |
| Mean EQ-5D* utility (Visit 1) | Treatment | 55.5 (SD: 19.4) | 62.7 (SD: 20.7) | 53.2 (SD: 31.8) | 50.6 (SD: 24.4) |
|  | Placebo | 50.1 (SD: 26.5) | 56.4 (SD: 22.5) | 50.5 (SD: 30.4) | 51.8 (SD: 30.6) |
| Mean EQ-5D* utility (Visit 2) | Treatment | 62.0 (SD: 21.2) | 62.1 (SD: 15.8) | NA |  |
|  | Placebo | 54.1 (SD: 23.0) | 57.5 (SD: 20.5) |  |  |
| Mean EQ-5D* utility (Visit 3) | Treatment | 65.3 (SD: 21.0) | 63.0 (SD: 16.7) | 64.6 (SD: 27.6) | 61.0 (SD: 26.7) |
|  | Placebo | 48.6 (SD: 24.4) | 56.6 (SD: 21.1) | 53.2 (SD: 32.3) | 52.5 (SD: 27.7) |

|  |  |  |  |  |
| --- | --- | --- | --- | --- |
| Patient comorbidities (n) | None | 29 (56%) | 25 (33%) | NA |
|  | Ataxia | 22 (42%) | 48 (63%) |  |
|  | Severe | 1 (2%) | 3 (4%) |  |
| Mean SFDs (Visit 1) | Treatment | 15.8 | 18.8 | NA |
|  | Placebo | 13.6 | 18.6 |  |
| Mean SFDs (Visit 2) | Treatment | 19.9 | 23.7 | NA |
|  | Placebo | 15.2 | 18.4 |  |
| Mean SFDs (Visit 3) | Treatment | 19.8 | 22.7 | NA |
|  | Placebo | 15.0 | 18.4 |  |

All patients maintained their existing standard of care and FFA or placebo was added
<sup>a</sup> Dosing in Study 1 was FFA 0.7mg/kg/day, up to a daily max of 26 mg/kg
<sup>b</sup> Dosing in Study 2 was FFA 0.4mg/kg/day, up to a daily max of 17 mg/kg
SD: Standard deviation; Mean SFD: Mean seizure free days in last 28-day period
\*EQ-5D values were multiplied by 100. For patients, PedSQL mapped to EQ-5D-Y, for carers EQ-5D-5L transformed to EQ-5D-3L

### Models of patient quality of life

The univariate analysis highlighted that patient age, 28-day frequency of SFDs, and having ataxia and severe motor impairments were statistically significant predictors of a patient’s QoL at baseline (Supplementary file 2, Table S1). The same qualitative patterns were found when PedsQL and EQ-5D-Y data at baseline were evaluated as the outcome variables (Supplementary File 2 Table S1). There were small quantitative changes in the coefficients estimated for each covariate in the PedsQL and EQ-5D-Y datasets. No interactions were statistically significant for either of the outcome datasets for patients (Supplementary Files 3 -6; Tables S2 – S5).

The reported patient QoL was highly variable between the data collection time points in the trials and between patients (Table 1, Supplementary file 9 Figure S1 and supplementary file 10 Figure S2). Therefore, subject ID and visit ID (visits 1, 2, 3) formed the two random effect components of the final mixed-effects model. Results of the final linear mixed-effects model for both outcome datasets for patients given the two random effects components are presented in Table 2.

**Table 2.** Final linear mixed-effects model results for 128 patients with EQ-5D-Y and PedsQL data.

| Covariate | Coefficients for EQ-5D-Y | Coefficients for PedSQL | p-value |
| --- | --- | --- | --- |
| 28-day frequency of SFDs | 0.55 | 0.274 | <0.001/<0.01 |
| Study 2 | 1.086 | 0.334 | >0.1/>0.05 |
| Age 6-11 years | 6.587 | -7.392 | >0.05/<0.001 |
| Age >12 years | 6.085 | -8.512 | >0.10/<0.001 |
| Motor impairments: Ataxia (relative to none) | -5.654 | - 2.962 | <0.05/<0.05 |
| Motor impairments: Severe<br>(relative to none) | -14.019 | -9.065 | >0.05/>0.1 |

### Linear mixed-effects model

As with the univariate analysis, similar qualitative patterns between EQ-5D-Y and PedsQL were observed in the final model covariates selected. One notable difference was that both age groups (Age 6-11 and Age >12 years old) had a statistically significant effect when analysing the PedsQL data but did not when assessing the EQ-5D-Y data. When adjusting for age and underlying comorbidities, results from the patient random effects regression model showed “frequency of SFDs per 28-days” as a significant predictor of QoL (gain in EQ-5D-Y utility of 0.005 per additional SFD, *p*<0.001), where the coefficient was divided by 100.

### Panel Linear Model

For the panel linear model 28-day frequency of SFDs was a statistically significant predictor of a patient’s QoL (Table 3).

**Table 3.** Final panel linear model results from 128 patients with EQ-5D-Y and PedsQL dataset.

| Covariate | Coefficient | Std. Error | p-value | Outcome variable |
| --- | --- | --- | --- | --- |
| 28-day frequency of SFDs | 0.660 | 0.220 | <0.01 | EQ-5D-Y |
| 28-day frequency of SFDs | 0.291 | 0.142 | <0.05 | PedsQL |

<sup>†</sup> For EQ-5D-Y coefficients refer to a 0-100 scale. All utility values predicted using these coefficients were divided by 100 before the predicted relationship was estimated (shown in Figure 1).

### Model selection

When comparing the mixed-effects regression model with the linear fixed effects panel regression model, the Hausman test *p*-value was >0.1, suggesting that the random-effects model was more appropriate for modelling the patient utility data (Table 3). Therefore, the mixed-effects models were selected as the final models for both patient datasets.

With the quantified and adjusted relationship between patient characteristics and patient EQ-5D-Y score calculated through the regression analysis, a patient’s utility score was predicted for each SFD (Figure 1) to visualise how a patient’s EQ-5D-Y score may vary with an increase in the number of SFDs.

**Figure 1.**
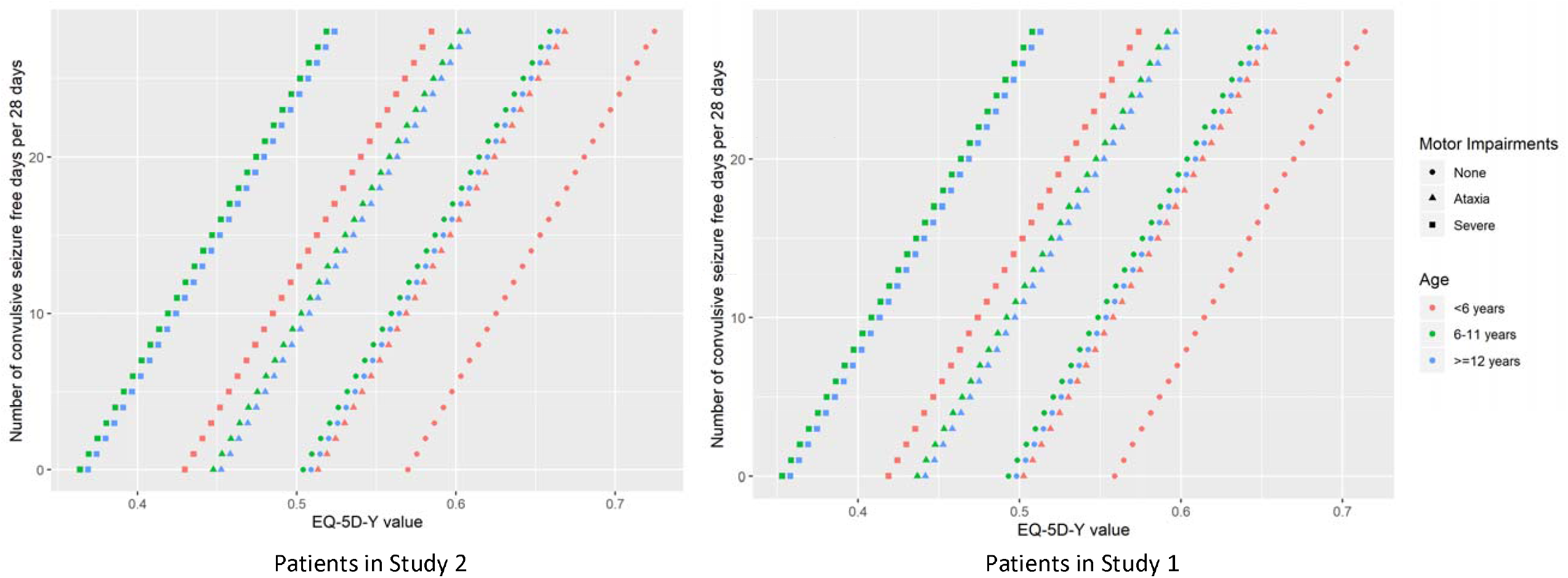
EQ-5D marginal means for patients who do (Study 2) and do not (Study 1) take concomitant stiripentol.

### Models of carer quality of life

The univariate analysis highlighted that age, 28-day frequency of SFDs at baseline were statistically significant predictors of carer QoL at baseline (Supplementary file 6, Table S5). Other than in the patient data, the patients’ motor impairments were not a significant predictor of carer QoL at baseline; however given the significance of motor impairments in the patient model and the possibility that motor impairments may be confounded by other covariates it was also tested in the selection of the final model. As with the patient models, none of the interactions between the covariates explored in the univariate analysis was statistically significant (p > 0.05) (Supplementary File 7, Table S6).

As with the patient data, the reported carer QoL varied between the two time points in the trials for which data were collected (Table 1) and between patients (Supplementary file 9 figure S1, supplementary file 10 figure S2 and supplementary file 11 figure S3). As per the patient model, subject ID and visit ID (visits 1, 2, and 3) formed the two random effect components of the final mixed effect model to account for this heterogeneity (Supplementary file 8, Table S7).

### Linear mixed-effects model

Results from the final mixed-effects model indicated that the 28-day frequency of SFDs and ataxia motor impairments were significant predictors of carer QoL (Table 4).

**Table 4.** Final mixed effect model results for EQ-5D-3L data 176 carers.

| Covariate | Coefficient | Std error | p-value |
| --- | --- | --- | --- |
| **28-day frequency of SFDs** | 0.513 | 0.227 | **<0.05** |
| **Study 2** | 3.069 | 3.755 | >0.1 |
| **Motor impairments – Ataxia**<br>(relative to none) | -27.597 | 8.281 | **<0.001** |
| **Motor impairments – severe**<br>(relative to none) | -13.659 | 9.483 | >0.1 |

<sup>†</sup> Coefficients refer to a 0-100 scale. All utility values predicted using these coefficients were divided by 100 before the predicted relationship was estimated (shown in Figure 1).

### Panel linear model

In the panel linear model, the only time-varying covariate representing the 28-day frequency of SFDs was evaluated. As with the patient models, and for the linear effects model, this covariate was a statistically significant predictor of a carer’s QoL. 28-day frequency of SFDs as a covariate was taken forward as the final covariate for the panel model (as previously described for the linear mixed-effects model).

### Model selection

Both the final panel linear model (Table 5) and final mixed-effects models (Table 4) were statistically compared using the Hausman test. The use of the linear panel model with fixed effects over the random effects model was statistically supported by the Hausman test (p <0.05) and was thus taken forward as the final model for the carer QoL data.

**Table 5.** Final panel linear model results for carer EQ-5D-3L data.

| Covariate | Coefficient | Std. Error | p-value |
| --- | --- | --- | --- |
| **28-day frequency of SFDs** | 1.361 | 0.386 | **<0.001** |

<sup>†</sup> Coefficients refer to a 0-100 scale. All utility values predicted using these coefficients were divided by 100 before the predicted relationship was estimated (shown in Figure 1).

The carer linear panel model showed “frequency of SFDs per 28-days” as a significant predictor of QoL (gain in EQ-5D-3L utility of 0.014 per additional SFD, *p*<0.001, where the coefficient was divided by 100).

As with the patient model, given the model coefficients estimated by the linear panel fixed effects model, the relationship was used to predict a carer utility score for each SFD that a patient experienced (Figure 2).

**Figure 2:**
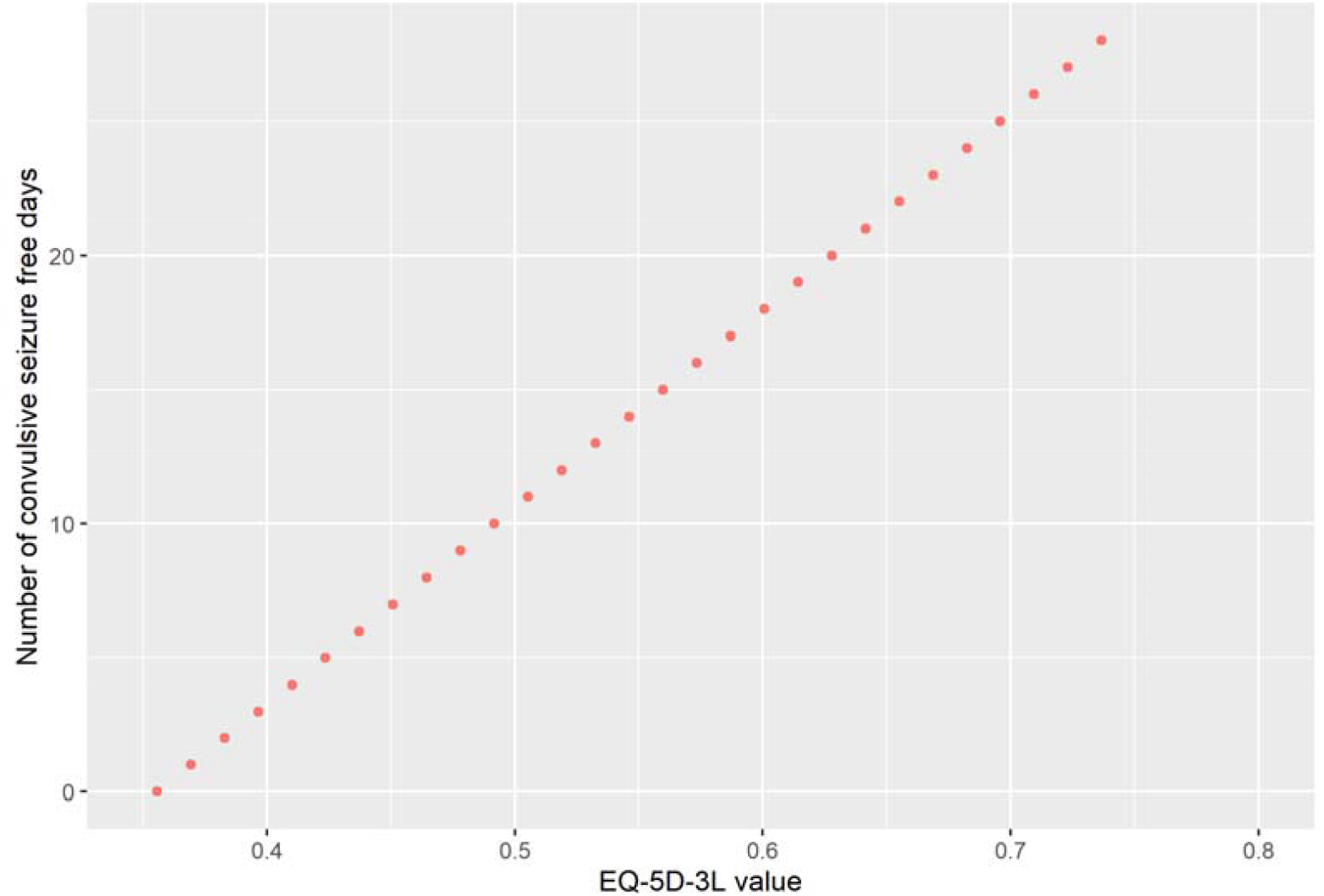
EQ-5D-3L marginal means for carers. *There was no separation by STP use as STP was not a significant covariate

## Discussion

The regression framework developed in this study identified key variables that impact the QoL of patients with DS and their carers, through the analysis of individual-level patient and carer data collected in the phase III registration trials for FFA. The results showed that the number of seizure free days (SFD) within a 28-day period had a significant impact on quality of life (QoL). This study provides quantitative evidence to support previous research indicating the positive impact of seizure free days on the QoL of both patients with DS and their carers (8,17,33). Age and comorbidities also affect patient QoL, supporting findings found in previous studies (34).

Previous studies that have explored the nature and impact of seizures in DS have focused on measurement of seizure frequency, or self-rated seizure severity, and used that to calculate the effect on QoL (15–17). However, a recent study (33) showed that DS impacts patients and carers beyond seizures, and highlighted the need for future clinical trials to fully explore the value of therapeutic interventions beyond simple seizure frequency reduction. It is also worth noting that whilst no studies identified in the SLR reported specific changes in utility values in relation to the number of SFDs, all five HTAs identified in this context modelled or evaluated a seizure-free health state (35–39). QoL data to inform cost-utility values within these studies were typically informed by data from Lennox Gastaut Syndrome, highlighting the need for studies to specifically evaluate data from DS patients and carers (see Supplementary file 1).

The method used in this study helps to understand the relative impact of treatment for DS, with incremental changes in SFDs contributing to incremental changes in QoL. It also demonstrates the importance of adjustment for important covariates when assessing QoL data. A typical DS patient aged <6 years with ataxia co-morbidities and 10 SFDs per 28-days would have an EQ-5D-Y utility of 0.56 and the carer would have an EQ-5D-3L utility of 0.49. If a patient’s SFDs increased to 20 or 28 (“seizure-free”) per 28 days (assuming <6 years of age and ataxia motor impairments), the patient utility would rise to 0.62 (+10.7%) or 0.66 (+19.2%), and their carer’s utility to 0.63 (+28.5%) or 0.73 (+48.9%), respectively. QoL of carers is impacted more by the frequency of SFD than that of patients. The highest achievable predicted carer QoL (e.g., no seizure days within a 28-day period) was 0.73, In comparison to average reported values in the UK population for adults of 0.857 (40), this is likely to have remained lower due to the burden of DS beyond seizures. This confirms the need to consider the impact of DS and DS treatments beyond seizures, and beyond the patient alone. An effective treatment not only has a direct health and QoL benefit to the patient but also has far reaching QoL benefit to the broader family unit affected by the patient’s condition.

There are several key strengths to the study. Firstly, the study utilised individual-level data prospectively collected at multiple points throughout clinical trials, favouring internal consistency and validity. Secondly, in general, similar qualitative patterns of statistical significance were seen for models of patient QoL using both outcome datasets (PedsQL and EQ-5D-Y) and using data from two separate studies. Although patient PedsQL data were mapped to EQ-5D-Y using the Khan mapping approach (22), which is reported to underestimate lower mapped utility values and could lead to potential underestimations, the results across the two patient outcome datasets were similar. Results were consistent across both sets of patient trial data, suggesting that the transformation of data is unlikely to have impacted the results. Furthermore, it is currently the only available approach to map PedsQL data to the more conventionally used EQ-5D typically used in health economics analyses. Thirdly, the regression analyses used two different analytical approaches in parallel to identify the most appropriate model for each dataset.

Despite a robust statistical approach, one limitation is that the analysis could only include data from two clinical trials with a small number of participants. Therefore, further work is needed to evaluate additional datasets to assess the replicability of our findings of an associating an increase in SFDs with an improved patient and carer QoL in DS; as well as the wider utility and generalisability of our approach. Given that individual-level data are routinely collected through clinical trials, encouraging the collection of patient and carer QoL data and analysis undertaken in this paper may help to further improve an understanding of which clinical and epidemiological factors have a quantifiable impact on QoL for both patients and carers. With an increasing need to understand the full patient/carer experience on treatment, and QoL being recognised as an important facet of treatment evaluation (41)(42), additional studies should be conducted to evaluate and quantify which clinical and epidemiological factors could lead to the biggest changes in QoL when DS patients are on treatment. Further research and economic evaluations should focus on evaluating the full range of health and quality of life effects on all members of the wider family, including siblings, who are affected by the patient’s condition.

## Conclusion

The current study showed there is a significant and quantifiable relationship between an increase in SFDs and a direct improvement in QoL for both patients and their carers. These results highlight the importance of increasing SFDs with effective antiseizure treatments, and the need for HTAs and other economic evaluations to consider the effectiveness of DS treatments beyond their impact on seizure reduction, and beyond the impact on patients alone.

## Data Availability

All data relevant to the study are included in the
online Supplementary Material. No additional
data are available.

