## Supplementary File for "Determining the Relationship between Seizure-Free Days and Other Predictors of Quality of Life in Patients with Dravet Syndrome and Their Carers"

**Table S1. Testing interaction terms following the univariate analysis for patient PedsQL.**

| **Covariate** | **Coefficient** | **p-value** | **Outcome variable** |
| --- | --- | --- | --- |
| Age*28-day frequency of seizure days at baseline | -0.005 | >0.1 | PedsQL baseline |
| Age*28-day frequency of SFDs at baseline | 0.005 | >0.1 | PedsQL baseline |
| Age * Study | -0.157 | >0.5 | PedsQL baseline |
| Age * Motor impairments | 0.775  0.913 | >0.1 | PedsQL baseline |
| 28-day frequency of seizure days at baseline *Study | 0.045 | >0.1 | PedsQL baseline |
| 28-day frequency of SFDs at baseline * Study | -0.045 | >0.1 | PedsQL baseline |
| 28-day frequency of seizure days at baseline * Motor impairments | 0.003  -0.128 | >0.1  >0.1 | PedsQL baseline |
| 28-day frequency of SFDs at baseline* Motor impairments | -0.003  0.128 | >0.1  >0.1 | PedsQL baseline |

**Table S2. Testing interaction terms following the univariate analysis for patient EQ-5D-Y data at baseline**

| **Covariate** | **Coefficient** | **p-value** | **Outcome variable** |
| --- | --- | --- | --- |
| Age* 28-day frequency of seizure days at baseline | 0.0271 | >0.1 | EQ-5D-Y baseline |
| Age* 28-day frequency of SFDs at baseline | -0.0271 | >0.1 | EQ-5D-Y baseline |
| Age * Study | -0.338 | >0.1 | EQ-5D-Y baseline |
| Age * Motor impairments | 1.238  1.736 | >0.1 | EQ-5D-Y baseline |
| 28-day frequency of seizure days at baseline * Study | 0.220 | >0.1 | EQ-5D-Y baseline |
| 28-day frequency of SFDs at baseline * Study | -0.220 | >0.1 | EQ-5D-Y baseline |
| 28-day frequency of seizure days at baseline * Motor impairments | 0.037  -0.031 | >0.1  >0.1 | EQ-5D-Y baseline |
| 28-day frequency of SFDs at baseline * Motor impairments | -0.037  0.031 | >0.1  >0.1 | EQ-5D-Y baseline |

^†^ For EQ-5D-Y coefficients refer to a 0-100 scale. All utility values predicted using these coefficients were divided by 100 before the predicted relationship was estimated (shown in Figure 1).

**Table S3. Stepwise testing of covariates with the random effects components of each model** **analysis for patient PedSQL and EQ-5D-Y data at the 3 follow-up time points**

| **Covariate** | **Coefficient** | **p-value** | **Outcome variable** |
| --- | --- | --- | --- |
| age + (1\|variable) + (1\|subjid) | -0.746 | **<0.001** | PedsQL |
| seizure day 28 freq + (1\|variable) + (1\|subjid) | -0.344 | **<0.001** | PedsQL |
| seizure free day 28 freq + (1\|variable) + (1\|subjid) | 0.344 | **<0.001** | PedsQL |
| Study + (1\|variable) + (1\|subjid) | 0.914 | >0.5 | PedsQL |
| physicalissues + (1\|variable) + (1\|subjid) | -4.098  -10.476 | **<0.05**  **<0.05** | PedsQL |
| age + (1\|variable) + (1\|subjid) | -0.767 | **<0.05** | EQ-5D-Y |
| seizure day 28 freq + (1\|variable) + (1\|subjid) | -0.647 | **<0.001** | EQ-5D-Y |
| seizure free day 28 freq + (1\|variable) + (1\|subjid) | 0.647 | **<0.001** | EQ-5D-Y |
| Study + (1\|variable) + (1\|subjid) | 1.409 | >0.1 | EQ-5D-Y |
| physicalissues + (1\|variable) + (1\|subjid) | -7.458  -15.333 | **<0.01**  **<0.05** | EQ-5D-Y |

^†^ For EQ-5D-Y coefficients refer to a 0-100 scale. All utility values predicted using these coefficients were divided by 100 before the predicted relationship was estimated (shown in Figure 1).

**Table S4. Testing interaction terms following the univariate analysis for carer EQ-5D-3L data at baseline**

| **Covariate** | **Coefficient** | **p-value** | **Outcome variable** |
| --- | --- | --- | --- |
| Age* 28-day frequency of seizure days at baseline | -0.130 | **<0.05** | EQ-5D-3L baseline |
| Age* 28-day frequency of SFDs at baseline | -0.130 | **<0.05** | EQ-5D-3L baseline |
| Age * Study | -1.451 | >0.1 | EQ-5D-3L baseline |
| Age * Motor impairments | 0.465  0.919 | >0.1 | EQ-5D-3L baseline |
| 28-day frequency of seizure days at baseline * Study | -0.268 | >0.1 | EQ-5D-3L baseline |
| 28-day frequency of SFDs at baseline * study | 0.268 | >0.1 | EQ-5D-3L baseline |
| 28-day frequency of seizure days at baseline * Motor impairments | 0.366  0.005 | >0.1  >0.1 | EQ-5D-3L baseline |
| 28-day frequency of SFDs at baseline * Motor impairments | -0.366  -0.005 | >0.1  >0.1 | EQ-5D-3L baseline |

^†^ Coefficients refer to a 0-100 scale. All utility values predicted using these coefficients were divided by 100 before the predicted relationship was estimated

**Table S5. Stepwise testing of covariates with the random effects components of each model** **analysis for carer EQ-5D-3L data at the 3 follow-up time points**

| **Covariate** | **Coefficient** | **p-value** | **Outcome variable** |
| --- | --- | --- | --- |
| age + (1\|variable) + (1\|subjid) | -0.728 | >0.05 | EQ-5D-3L |
| seizure day 28 freq + (1\|variable) + (1\|subjid) | -0.473 | **<0.05** | EQ-5D-3L |
| seizure free day 28 freq + (1\|variable) + (1\|subjid) | 0.473 | **<0.05** | EQ-5D-3L |
| Study + (1\|variable) + (1\|subjid) | 1.770 | >0.1 | EQ-5D-3L |
| physicalissues + (1\|variable) + (1\|subjid) | -26.821  -14.998 | **<0.001**  >0.1 | EQ-5D-3L |

^†^ Coefficients refer to a 0-100 scale. All utility values predicted using these coefficients were divided by 100 before the predicted relationship was estimated

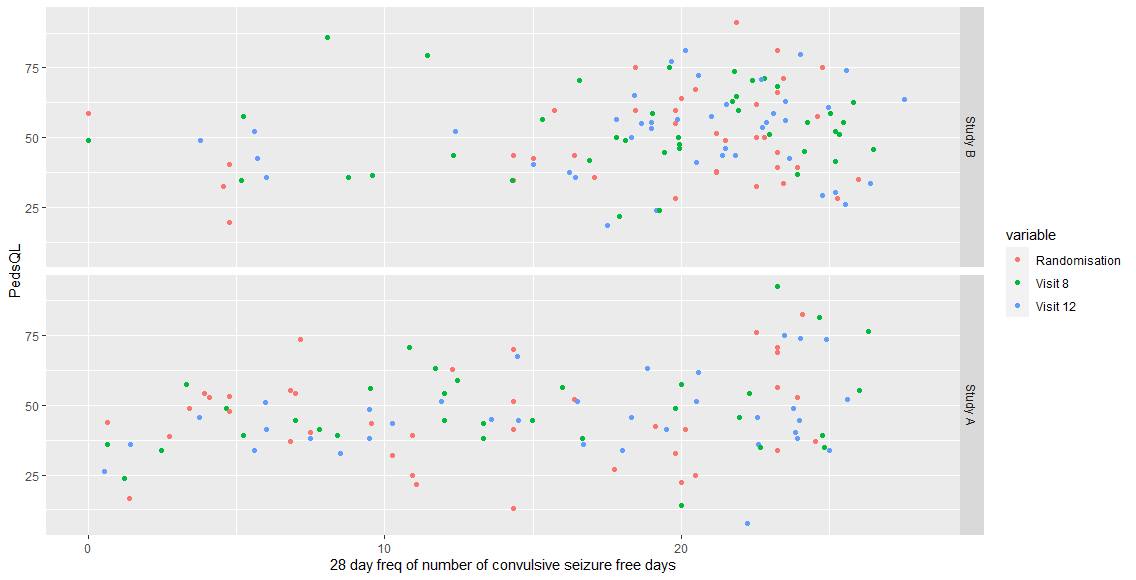
**Figure S1. PedsQL data for 128 patients in both study groups of the trial for each follow-up point.**

**Figure S2. EQ-5D-Y data for 128 patients in both study groups of the trial for each follow-up point.**

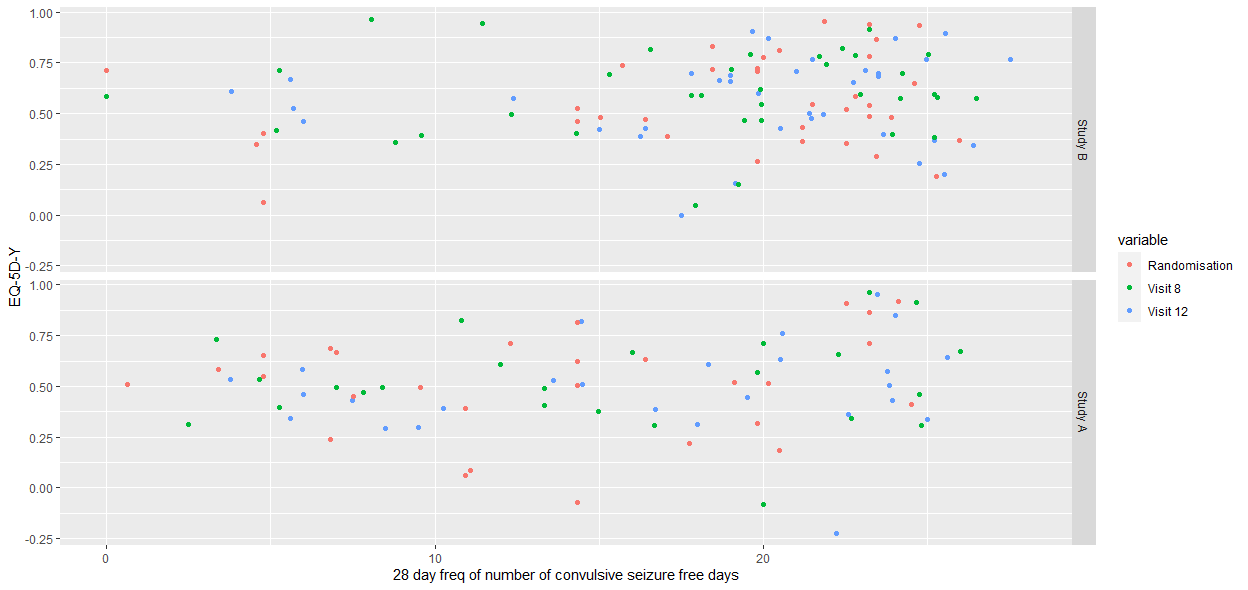

**Figure S3. EQ-5D-3L data for 176 carers in both study groups of the trial for each follow-up point.**

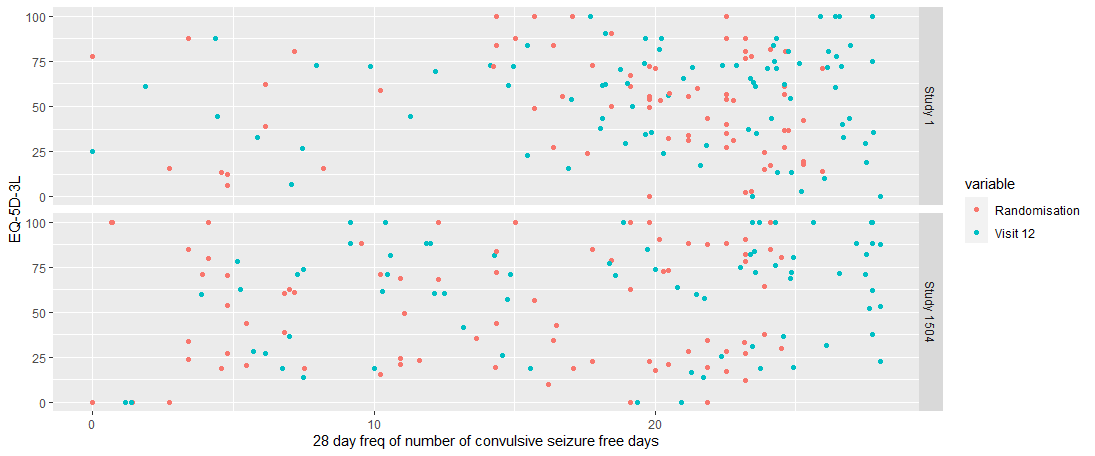
